# Identification and Estimation of Undetected COVID-19 Cases Using Testing Data from Iceland

**DOI:** 10.1101/2020.04.06.20055582

**Authors:** Karl M. Aspelund, Michael Droste, James H. Stock, Christopher D. Walker

## Abstract

In the early stages of the COVID-19 pandemic, international testing efforts tended to target individuals whose symptoms and/or jobs placed them at a high presumed risk of infection. Testing regimes of this sort potentially result in a high proportion of cases going undetected. Quantifying this parameter, which we refer to as the *undetected rate*, is an important contribution to the analysis of the spread of the SARS-CoV-2 virus. We show that partial identification techniques can credibly deal with the data problems that common COVID-19 testing programs induce (i.e. excluding quarantined individuals from testing and low participation in random screening programs). We use public data from two Icelandic testing regimes during the first month of the outbreak and estimate an identified interval for the undetected rate. Our main approach estimates that the undetected rate was between 89% and 93% before the medical system broadened its eligibility criteria and between 80% and 90% after.

## 1 Introduction

Estimates of viral prevalence in the early stages of a pandemic are of vital importance because of their influence on the timing and stringency of public health interventions. In the first month of the COVID-19 outbreak — and, in many regions of the United States, even today — eligibility for testing was restricted to individuals thought to be at the highest risk of a positive diagnosis. For instance, in most of the U.S., testing was initially restricted to individuals presenting severe flu-like symptoms or a known contact with a confirmed infection.^1^ Narrow testing criteria lead to the possibility that a large share of the infected population are not eligible for testing and many cases go undetected by public health authorities. Consequently, health authorities have limited understanding of viral prevalence in the early stages of the pandemic. This concern is just as important as social distancing measures are lifted in many countries.

Our paper attempts to identify and estimate the share of infections that go undetected under these testing guidelines, a parameter that we call *the undetected rate*. We consider COVID-19 testing in Iceland because the operation of two distinct testing regimes provide valuable identifying information. The first testing regime, operated through the Icelandic medical system, targeted individuals with high presumed risk of infection with eligibility guidelines that closely resembled those in the United States and other developed countries. A second testing program was a voluntary program ran by deCODE Genetics, a biopharmaceutical company, that non-quarantined Icelanders qualified for. The firm also ran a separate random screening experiment on the Icelandic population. Both programs started within two weeks of Iceland’s first confirmed infection.

Although deCODE Genetics testing data is informative about the ineligible subpopulation, it is insufficient to credibly point identify the undetected rate. Indeed, deCODE testing is voluntary, excludes quarantined individuals, and the random screening suffered from relatively low participation (approximately 34%). Motivated by these concerns, we derive two types of bounds for the undetected rate and estimate them using publicly available COVID-19 testing data and results published by Guðbjartsson et al. (2020*b*). All results are from testing before the peak caseload of confirmed active infections was reached on April 5.^2^ We also discuss the construction of a least-favorable confidence set for the bounds that overcomes limitations of COVID-19 testing data. In a related work, Manski and Molinari (2020) partially identify COVID-19 infection rates in the United States and Italy.^3^

Across the approaches and over different time periods, we estimate a high undetected rate. One approach uses data from voluntary deCODE testing, and accounts for the exclusion of quarantined individuals and the over-representation of symptomatic individuals in the sample. We estimate bounds on the undetected rate of 89% to 93% before the medical system widened its eligibility criteria and 80% to 90% after. This approach relies on estimation of the share of the non-infected who are eligible, using historical data on flu symptom prevalence. A second approach avoids this parameter and uses deCODE’s randomized encouragement design, estimating wider bounds from 68% to 92%. The width of the bounds reflects difficulty in identifying the rate of infection for those who did not participate when (randomly) invited. More direct random testing methods that do not exclude quarantined individuals are necessary if researchers want to estimate pandemic parameters in an informative way. Overall, the results speak to the need for testing policies with broad criteria extending well beyond individuals deemed to be at high risk.

Our efforts complement other studies on the rate of undetected COVID-19 cases. Many studies estimate the infection rate in the population and compare these estimates to positive testing rates from existing testing programs to suggest large fractions of undetected or asymptomatic infections (Li et al., 2020; Mizumoto et al., 2020; Nishiura et al., 2020; Russell et al., 2020; Wu et al., 2020*a*; Qiu, 2020). In recent weeks, results from serological testing were used for a similar exercise (New York Governor’s Press Office, 2020; Public Health England, 2020; Spanish Ministry of Health, 2020; Wu et al., 2020*b*; Boston Public Health Commission, 2020; Public Health Agency of Sweden, 2020; Streeck et al., 2020); comparisons with the rate of positive test cases suggest a large fraction of cases went undetected (Vogel, 2020, e.g.).

Our goal is more specific: we seek to evaluate the detection rate of testing protocols that focus on the highly symptomatic and high-risk groups, the kind of criteria that prevailed early in the pandemic in particular. The advantage of the Icelandic setting is that we are able to undertake such a comparison in a single population and at different points in time as the virus spread, due to the presence of different testing programs using the same methods but of varying eligibility criteria. Moreover, the identification challenges emphasize that careful test protocol design—e.g. broad eligibility (including for those in quarantine), randomization approaches that ensure high participation—are vital to allow accurate and simple analysis of viral spread. If testing efforts exclude certain groups or are mostly voluntary despite randomized encouragement—common elements of the serological studies cited above and other swab-based testing efforts (Office for National Statistics, 2020; Indiana State Dept. of Health, 2020, e.g.)—then researchers can rigorously analyze the results of testing programs using our methods. The inference problems speak to an important opportunity for econometricians in the study of pandemics.

Section 2 summarizes the Icelandic testing efforts in the first month of the pandemic. Section 3 outlines the methods we use to identify the undetected rate. Section 4 summarizes our estimation procedure and inference. Section 5 reviews the data and summarizes the results.

## 2 COVID-19 Testing in Iceland

There have been two separate testing regimes operating in Iceland since the middle of March 2020, approximately two weeks after the first confirmed infection in Iceland. Guðbjartsson et al. (2020*b*) describes both of these testing regimes in detail.

The first testing program is through the Icelandic healthcare system, which tested individuals exhibiting severe symptoms (cough, fever, muscle ache, and shortness of breath) and/or at high risk of infection because of close contact with a diagnosed individual or travel in “high-risk areas.” On March 19, the Directorate of Health broadened the criteria to include anyone returning from abroad and any symptomatic individual.^4^ Samples are taken at local clinics or the National University Hospital of Iceland (NUHI) and analyzed by the hospital’s microbiology department. We refer to this as NUHI testing or medical testing.

The second is a free, voluntary program administered by the biopharmaceutical company deCODE Genetics. The deCODE program began on March 13 and is open to individuals who have no or few symptoms and have not been tested through NUHI. Later, deCODE invited a random sample of Icelanders for a COVID-19 test over text message. There were no restrictions based on previous testing or symptoms. Testing of the random sample began on April 1, and by April 4, 33.7% of those invited had participated. NUHI and deCODE used the same methods to extract samples and the viral RNA and used similar methods to detect the presence of the virus (Guðbjartsson et al., 2020*b*).

Notably, quarantined individuals were not permitted to show up for deCODE testing, even if they were invited to participate in random screening. Individuals are placed under quarantine if they have been in close contact with a diagnosed individual or have been traveling, initially in high-risk areas and later anywhere abroad.^5^ The share of the population under quarantine went up to almost 3% in the first month of the COVID-19 outbreak.

## 3 Identification

This paper attempts to estimate the rate of COVID-19 infection that is undetected by the NUHI testing program. A prerequisite for statistical inference is determining the identification region for the parameter of interest and in this section we study that problem. Formally, we would like to estimate *θ* ≡ Pr(*E* = 0|*I* = 1), where the random variables *E ∈ {*0, 1*}* and *I ∈ {*0, 1*}* indicate NUHI test eligibility and COVID-19 infection, respectively. The parameter represents NUHI test ineligibility rate for the infected sub-population and is referred to as the *undetected rate*.

Several aspects of the data complicate identification of *θ*. To see this, Bayes rule and the law of total probability yield

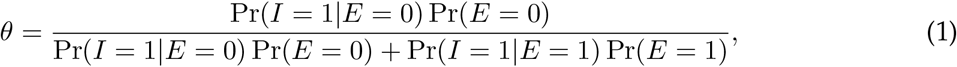

which implies that if Pr(*E* = 1), Pr(*I* = 1|*E* = 0), and Pr(*I* = 1|*E* = 1) are known, then *θ* is point identified. A naïve approach to identifying Pr(*I* = 1|*E* = 0) is to use the positive test rate for deCODE volunteers.^6^ Recognizing selection issues associated with volunteerism and that deCODE testing excludes quarantined individuals, such an approach is implausible. Consequently, we propose two alternative identification strategies: one that avoids Pr(*I* = 1|*E* = 0) altogether and another that partially identifies it. We will refer to these approaches as the *random screening method* and *odds ratio method*, respectively, and describe each formally in the following subsections.

Both methods of identifying *θ* require identification of Pr(*I* = 1|*E* = 1) and Pr(*E* = 1). Let *T ∈ {*0, 1*}* be a random variable that indicates participation in NUHI testing. Challenges for identification arise because *{E* = 1*}* = *{E* = 1, *T* = 1*}* ∪ *{E* = 1, *T* = 0*}* and *{E* = 1, *T* = 0*}* cannot be studied using the data in an informative way. The next assumption addresses this problem and is maintained throughout the paper.

**Assumption 1.** *The event {T* = 1*} is equivalent to {E* = 1*}*.

Assumption 1 states that all NUHI-eligible individuals are tested and therefore *{E* = 1, *T* = 0*}* = *∅*. In practical terms, Pr(*E* = 1) and Pr(*I* = 1|*E* = 1) are identified by Pr(*T* = 1) and Pr(*I* = 1|*T* = 1), respectively. By construction *{T* = 1*}* implies *{E* = 1*}*, so the credibility of the assumption rests on whether *{E* = 1*}* implies *{T* = 1*}*. We believe this is reasonable because the severity of the symptoms characterizing NUHI eligibility and the heightened salience of symptoms due to the pandemic mean that it is likely that all eligible individuals reported for testing.^7^ In line with Assumption 1, we maintain the *E* notation throughout the paper rather than substituting *T*.

### 3.1 Random Screening

The randomized encouragement design provides a method for partially identifying the undetected rate *θ*. First, an application of Bayes rule and the law of total probability yields an expression for the undetected rate,

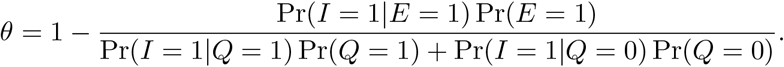

where *Q ∈ {*0, 1*}* is a random variable that indicates quarantine status. Next, we note the following:

1. The design is uninformative about the distribution of *I*|*Q* = 1 because those in quarantine are ineligible for deCODE testing. Consequently, Pr(*I* = 1|*Q* = 1) *∈* [0, 1] without further assumptions.
2. Due to participation issues, the distribution of *I*|*Q* = 0 is partially identified. The data is uninformative about distribution of *I*|*Q* = 0, *Y* = 0, where *Y ∈ {*0, 1*}* is a random variable that indicates whether an individual participated in the testing after being invited. The lack of information on the infection rate of non-participants and the law of total probability leads to Pr(*I* = 1|*Q* = 0) *∈* [*L*_0_, *U*_0_], where

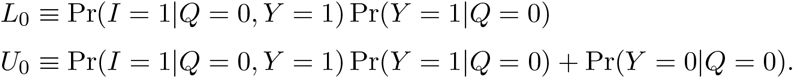

These observations imply 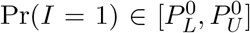, where 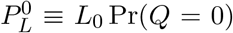 and 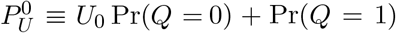. Immediately, an identification region for *θ* is generated and is summarized in Proposition 1.

#### **Proposition 1**.

*Under the randomized encouragement design and Assumption 1*, 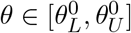 *where*

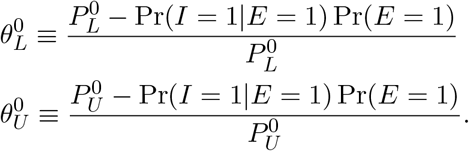

*Proof*. See Appendix Section A.

We introduce further distributional assumptions to sharpen inference about *θ*.

**Assumption 2**. *The joint distribution of the data satisfies 1*. Pr(*I* = 1|*Q* = 0) ≤ Pr(*I* = 1|*Q* = 1), *2*. Pr(*I* = 1|*Q* = 1) ≤ Pr(*I* = 1|*E* = 1), *and 3*. Pr(*I* = 1|*Q* = 0, *Y* = 0) ≤ Pr(*I* = 1|*Q* = 0, *Y* = 1)

The first part of Assumption 2 is a monotonicity restriction that states that those in quarantine are more likely to be infected than those not in quarantine. The restriction is plausible because a person is placed in quarantine if they have been in contact with an infected individual and/or have travelled from a high-risk area. The second part of Assumption 2 states that the NUHI infection rate is higher than the quarantine infection rate and is reasonable because of the emphasis on symptoms for NUHI testing whereas asymptomatic individuals can be quarantined. Finally, Part 3 is a monotonicity restriction that imposes that the infection rate of participants is higher than that of nonparticipants. In other words, those who are at higher risk of infection may be more concerned about their infection status and more likely to participate when invited for testing.

Assumption 2 tightens the identified set for *θ* by narrowing the bounds on the infection rate. Indeed, the assumption implies that Pr(*I* = 1|*Q* = 1) *∈* [Pr(*I* = 1|*Q* = 0), Pr(*I* = 1|*E* = 1)] and therefore 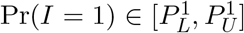, where 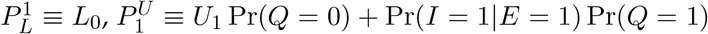,and *U*_1_ = Pr(*I* = 1|*Q* = 0, *Y* = 1). Proposition 2 formalizes the previous discussion.

#### **Proposition 2**.

*Under Assumption 2*, 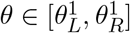 *where*

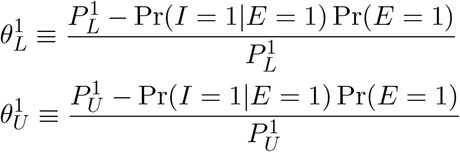

*Proof*. See Appendix Section A.

**Remark 1**. The driver of the identification results are bounds for Pr(*I* = 1) that use the information embedded in the randomized encouragement design. Manski and Molinari (2020) derive bounds for Pr(*I* = 1) in the context of COVID-19 testing in Illinois, New York, and Italy under different assumptions. We report their bounds using Icelandic data in Appendix Section C.

### 3.2 Odds Ratio

We now present an alternative approach to partially identifying *θ* that does not make use the randomized encouragement design. We call this approach the *odds ratio method* and the label arises because one can write

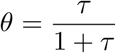

where

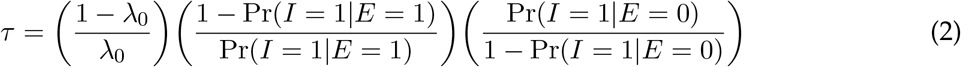

and *λ*_0_ ≡ Pr(*E* = 1|*I* = 0). A lemma in Appendix Section A establishes this representation. The parameter *λ*_0_ is interpreted as the baseline rate of cough/cold/flu symptoms that are severe enough to qualify for a NUHI test, even though the individual is not infected. It is calibrated using data from past flu seasons. Every year, the Health Directorate receives weekly counts of all visitors to primary-care centers and health clinics with flu-like symptoms, regardless of the purpose of the visit.^8^ They are not necessarily tested for influenza. In particular, we use the highest weekly caseload across the flu seasons from 2009 through mid-2017 (the latest year available) as a proportion of the population. This occurred in November 2009 during the H1N1 flu pandemic, when in one week 2,012 individuals visited health clinics with flu-like symptoms, or 0.63% of the population.

Under Assumption 1 and the expression (2), we only need to identify the conditional distribution of *I* given *E* = 0. To this end, let *C ∈ {*0, 1*}* and *D ∈ {*0, 1*}* be random variables equal to 1 if an individual is symptomatic and deCODE volunteer, respectively. We make the following assumption to derive bounds for Pr(*I* = 1|*E* = 0).

**Assumption 3**. *The following is true: 1*. Pr(*C* = 1|*Q* = 0, *E* = 0) *∈* [*λ*_0_, Pr(*C* = 1|*D* = 1)], *and 2*. Pr(*I* = 1|*E* = 0, *Q* = 1) ≤ Pr(*I* = 1|*E* = 1).

Part 1 of Assumption 3 states that the symptom rate for the unquarantined, NUHI-ineligible subpopulation is no less than the baseline cold/cough/flu symptom rate and does not exceed the symptom rate for deCODE volunteers. Because deCODE testing is voluntary, those with COVID-like symptoms will be more concerned about their infection status and select into testing. If so, the symptom rate of deCODE volunteers is an upper bound on the symptom rate as a whole. This intuition is similar to Part 3 of Assumption 2. Meanwhile, the symptom rate must at least the rate of severe flu-like symptoms, so the lower bound is *λ*_0_. Part 2 states that the infection rate of the NUHI-ineligible, quarantined individuals is less than the NUHI eligible infection rate. The plausibility of this assumption follows a similar argument to Part 2 of Assumption 2.

Now that we have made this assumption, we work through the identification of Pr(*I* = 1|*E* = 0). By the law of total probability, we can decompose Pr(*I* = 1|*E* = 0) as follows

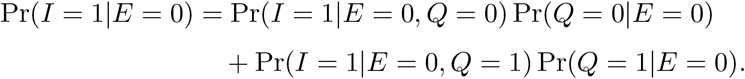

We consider each term individually:

1. The law of total probability yields

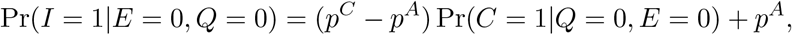

where *p*^*A*^ ≡ Pr(*I* = 1|*E* = 0, *Q* = 0, *C* = 0) and *p*^*C*^ ≡ Pr(*I* = 1|*E* = 0, *Q* = 0, *C* = 1). This and Part 1 of Assumption 3 yields Pr(*I* = 1|*E* = 0, *Q* = 0) *∈* [*f* (*λ*_0_), *f* Pr(*C* = 1|*D* = 1)], where *f* (*x*) = (*p*^*C*^ − *p*^*A*^)*x* + *p*^*A*^ for any *x ∈* [0, 1].
2. Part 2 of Assumption 3 yields Pr(*I* = 1|*E* = 0, *Q* = 1) *∈* [0, Pr(*I* = 1|*E* = 1)].
3. Bayes rule and Pr(*E* = 0|*Q* = 1) *∈* [0, 1] yields Pr(*Q* = 1|*E* = 0) *∈* [0, Pr(*Q* = 1)*/* Pr(*E* = 0)].

These observations imply an identified set for Pr(*I* = 1|*E* = 0) given by

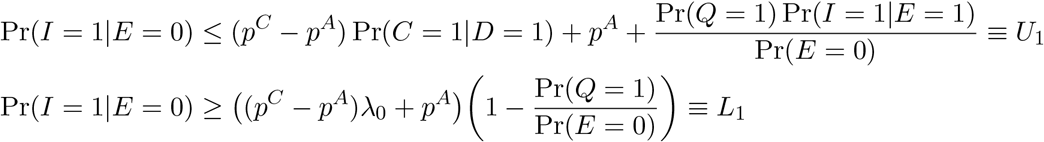

Let *τ*_*U*_ and *τ*_*L*_ denote *τ* evaluated at *U*_1_ and *L*_1_, respectively. The next proposition states the immediate identified set for *θ*.

#### **Proposition 3**.

*Assumptions 1 and 3 imply that* 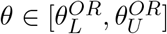, *where*

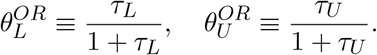

*Proof*. See Appendix Section A.

### 3.3 Sensitivity and Specificity

Our identification has implicitly assumed the absence of false positives and false negatives in tests administered by both the NUHI and deCODE testing regimes. Formally, we are assuming that Pr(*I* = 1|*A*) = Pr(*R* = 1|*A*), where *R ∈ {*0, 1*}* indicates a positive test and *A* is some conditioning event (for example, *A* = *{E* = 1*}*). The distinction is important because

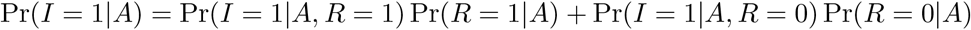

by the law of total probability, which means that without further assumptions the infection rate is an affine function of the positive test rate. Identification of the components of Pr (*I* = 1|*A*) are discussed in Manski and Molinari (2020) and, of course, a limitation of our approach is that we are not agnostic about this parameter.

However, there is reason to believe that our specificity and sensitivity assumptions are plausible. Both testing regimes use reverse transcriptase polymerase chain reaction (RT-PCR) tests with specimens collected via nasopharyngeal swabs (Guðbjartsson et al., 2020*b*). This testing system has been adopted by the vast majority of developed countries and is considered to be the ‘gold standard’ of viral testing (Ramdas, Darzi and Jain, 2020). Analytical estimates of test sensitivity and specificity, which are provided by RT-PCR test manufacturers by analyzing samples with known results, suggest that test sensitivity and specificity under ideal conditions are both close to 100%.^9^ False negatives are most likely to occur when viral load is low — a cycle threshold value below 35 or 40, depending on sampling method (Bullard et al., 2020; Zheng et al., 2020) — e.g. shortly after exposure, when it is not clear that infected individuals are able to infect others. For our purposes, the ability to infect others, rather than viral presence alone, should determine infected state *I*, and therefore we consider the ability to detect truly infectious individuals to be quite high.

While we believe that our baseline estimates are reasonable given the evidence on RT-PCR test sensitivity and specificity, an extension that explicitly accounts for these factors would likely be important in applying our methods to alternative testing regimes; for instance, in the study of ongoing antibody tests that are believed to exhibit lower test sensitivity and specificity.

## 4 Estimation and Inference

The bounds derived in the previous section are simple to compute. We know the population counts for some variables, meaning that we can measure some parameter with certainty. For example, Pr(*Q* = 1) = *N*_*Q*_*/N* where *N*_*Q*_ is the number of individuals in quarantine and *N* is the Icelandic population. All the parameters in the odds ratio approach can be computed this way.

The data for the random screening approach is based on a random sample of the Icelandic population. Consequently, some parameters need to be estimated and in such case we use sample counterparts. For example, we do not observe population counts of individuals with *Q* = 0 and *Y* = 1, but rather members of the subpopulation who appear in the random sample of invitees in the random screening approach.

The only nonstandard aspect of the estimation procedure relates to Pr(*I* = 1, *E* = 1). Despite the plausibility of Assumption 1, it is possible that in practice there is a lag between becoming eligible for and receiving a NUHI test. This motivates the *eligibility window method* for computing the joint probability mass. Let 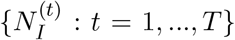 be a time series, where 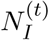 is the NUHI positive test count for day *t ∈ {*1, …, *T}* and *T* is a user-defined number of days (a tuning parameter). The eligibility window method computes Pr(*I* = 1, *E* = 1) using 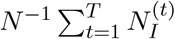. While it is unrealistic to assume that all eligible individuals report for NUHI testing on a single day, we find it reasonable to assume that across a window of days the medical system comes close to testing all those who become eligible. Therefore the sum ameliorates the stock-flow problem.

We now turn to the problem of constructing a confidence region for the identified set under the random screening approach. The main takeaway is that it is hard to make powerful inferences about pandemic parameters with limited public health data. Consequently, there is an important role for rigorous data collection methods (i.e. monitored quarantine, contact tracing) to aid the statistical analysis of pandemics. It must be noted that the focus of this paper is identification; the following discussion is to point out another inferential challenge in the econometric analysis of pandemics and serves to motivate future research in this area.

Let **P** be the joint distribution of the observed data and assume that **P** *∈ 𝒫*, where *𝒫* is a family of distributions. The identified set under **P**, denoted Θ_*I*_(**P**), is a compact subset [*θ*_*L*_, *θ*_*U*_] ⊆ [0, 1]. In the spirit of Horowitz and Manski (2000), a candidate 1 − *α* confidence set for Θ_*I*_(**P**) is given by

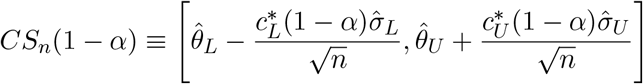

where 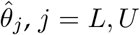 are estimators for the bounds, 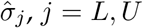 estimate their standard deviations, and *n* is the sample size. The critical values are defined as

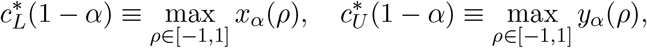

Where

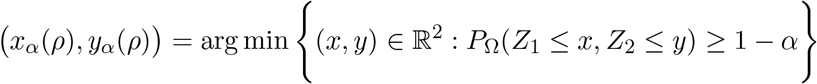

for each *ρ ∈* [−1, 1], and *P*_Ω_(*·, ·*) is the distribution function for bivariate normally distributed random vector with mean 0_2_ and correlation matrix Ω. See Appendix Section A for high-level assumptions for the confidence set to satisfy

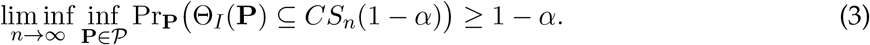

The proposed critical values are unconventional. The construction borrows from the analytical example in Cocci and Plagborg-Møller (2019) and ultimately reflects data availability. Indeed, the bounds are nonlinear functionals of **P** so deriving a closed-form expression for 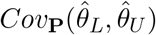 is difficult. Moreover, the lack of individual-level data precludes the use of the nonparametric bootstrap/subsampling as a sensible way to perform inference on the bounds. The optimization means that the critical values reflect the least-favorable correlation structure. Consequently, the confidence set may be conservative (i.e. (3) holds strictly).

A similar issue is embedded in the computation of 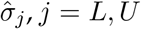. A standard approach for computing 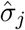 is the delta method, but limitations of publicly available COVID-19 data means that some off-diagonal elements of the variance matrix under **P** are nonidentified.^10^ Since our data is comprised of indicator variables, covariances lie in a compact interval meaning that worst-case estimates of 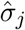 can be obtained by maximization. Indeed, we can compute the variance implied by the delta method as a function of the nonidentified parameters and then maximize over their feasible values.^11^ Such approach adds an additional layer of conservatism to the inference and motivates future econometric research in the area of powerful inference under pandemic data limitations.

## 5 Empirical Analysis

### 5.1 Data

We use data released by Icelandic public health authorities on the COVID-19 pandemic and on normal flu seasons. The Icelandic Directorate of Health releases daily counts of new confirmed cases and tests conducted on its COVID-19 information website.^12^ We use these data where possible. However, infection rates for individuals with no vs. mild symptoms are only available in the results from Guðbjartsson et al. (2020*a*), who report infection and test counts by symptom status for voluntary deCODE testing between March 13 and 19. Results from the randomized screening are also available in Guðbjartsson et al., who report infection and test counts for the random sample tested between April 1 and 4. Only the most updated quarantine counts are available on the COVID-19 website, so we use daily announcements by the Icelandic Department of Civil Protection and Emergency Management to find past quarantine totals as needed for calculating bounds.

To estimate *λ*_0_ in the odds ratio method, we use data from the Directorate of Health’s website with weekly counts of individuals who come to primary-care center or health clinics with flu-like symptoms for every flu season from the 2009-10 through the 2017-18 flu season (the most recent season available).^13^ Appendix Section D contains the precise details on the data and construction of each parameter value. Table 1 summarizes the estimates used to construct the bounds.

**Table 1:**
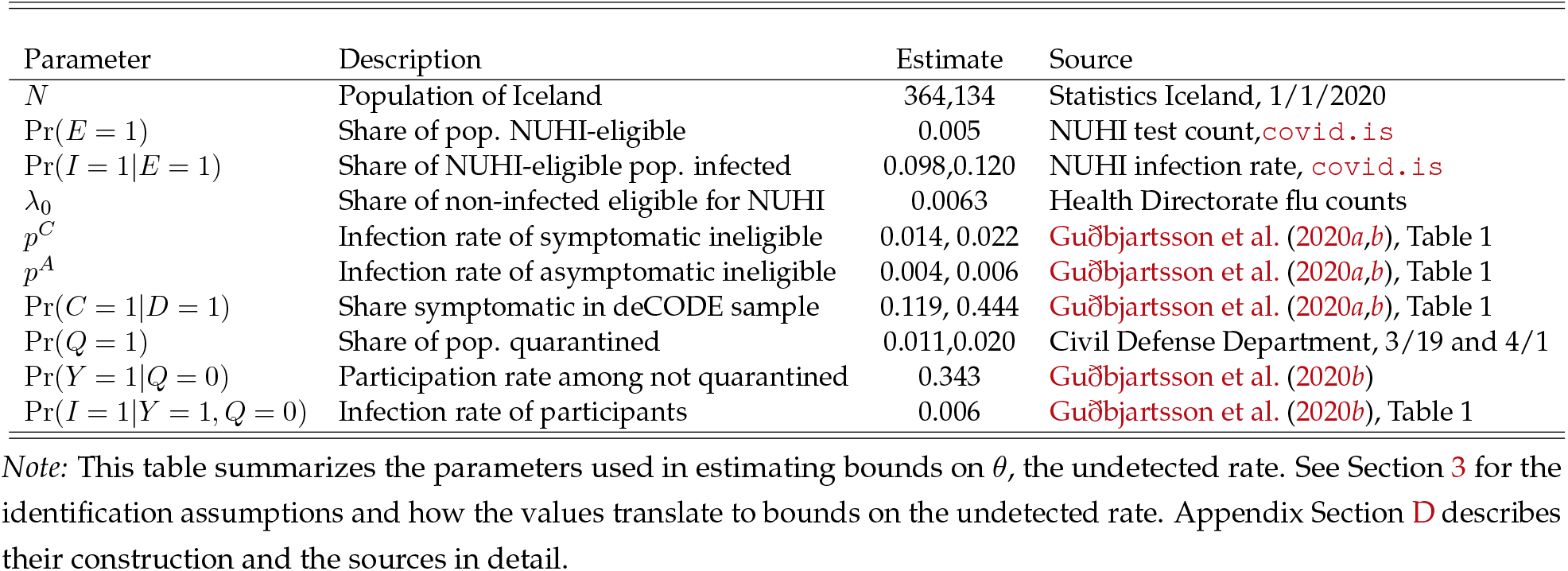
Parameter Values and Data Sources.

### 5.2 Results

The results are summarized in Table 2. While our approaches rely on different assumptions, the results are consistent: more than two thirds of all infected Icelanders are ineligible for testing in the medical system, even after testing criteria were broadened. The results rely on a series of important assumptions on NUHI eligibility and lags in testing (see Assumption 1 and Section 4) and the plausibility of deCODE results as estimates of the ineligible rate, which motivated our use of partial identification techniques. We explore the results in detail below.

**Table 2:** Estimates of the Proportion NUHI-Ineligible among the Infected.

| Method | Undetected Rate $\theta$ | | Dates of<br>NUHI Testing | Dates of<br>deCODE Testing |
| --- | --- | --- | --- | --- |
|  | Lower Bound | Upper Bound |  |  |
| Random Screening<sup>a</sup> | 0.675 | 0.999 | 4/1 - 4/4 | 4/1 - 4/4 |
| Random Screening<sup>b</sup> | 0.681 | 0.922 | 4/1 - 4/4 | 4/1 - 4/4 |
| Odds Ratio | 0.891 | 0.938 | 3/13 - 3/19 | 3/13 - 3/19 |
| Odds Ratio | 0.806 | 0.905 | 4/1 - 4/4 | 4/1 - 4/4 |

Notes: This table summarizes the methods we outline to estimate $\theta = P(E = 0|I = 1)$ , the undetected rate. Section 3 describes the methods in detail. The random screening method relies on test results from a sample of Icelanders randomly invited to participate in deCODE testing from April 1 through April 4. The odds ratio method relies on the parameter $\lambda_0 = P(E = 1|I = 0)$ , which we estimate based on data from past flu seasons, as well as infection rates of the symptomatic and asymptomatic from Guðbjartsson et al. (2020b). Parameter values and their construction are summarized in Table 1 and described in detail in Appendix Section D.
<sup>a</sup> Relies only on Assumption 1.
<sup>b</sup> Relies on Assumptions 1 and 2.

**Table 3:** Estimation of Population Infection Rate following Manski and Molinari (2020)

| Date | Lower Bound | Upper Bound |
| --- | --- | --- |
| 3/13 | 0.0001475 | 0.4084 |
| 3/14 | 0.0001480 | 0.4084 |
| 3/15 | 0.0003447 | 0.4084 |
| 3/16 | 0.0005281 | 0.4084 |
| 3/17 | 0.0005281 | 0.4092 |
| 3/18 | 0.0006923 | 0.4092 |
| 3/19 | 0.0006923 | 0.4092 |
| 3/20 | 0.0006923 | 0.4092 |
| 3/21 | 0.0006923 | 0.4092 |
| 3/22 | 0.0006923 | 0.4092 |
| 3/23 | 0.0006923 | 0.4092 |
| 3/24 | 0.0006923 | 0.4092 |
| 3/25 | 0.0006923 | 0.4092 |
| 3/26 | 0.0006923 | 0.4092 |
| 3/27 | 0.0006923 | 0.4092 |
| 3/28 | 0.0006923 | 0.4092 |
| 3/29 | 0.0006923 | 0.4092 |
| 3/30 | 0.0006923 | 0.4092 |
| 3/31 | 0.0006923 | 0.4092 |
| 4/1 | 0.0006923 | 0.4092 |
| 4/2 | 0.0006923 | 0.4092 |
| 4/3 | 0.0006923 | 0.4092 |
| 4/4 | 0.0006923 | 0.4092 |

Notes: The table shows daily bounds on the population infection rate following Equation (13) of Manski and Molinari (2020). See Remark 1 and Appendix Section C for details.

#### Odds ratio method

Since this method relies on deCODE infection rates by symptom status when bounding, we limited to two time periods for which we have such data from Guðbjartsson et al. (2020*b*), i.e. March 13 through 19—available in the pre-print (Guðbjartsson et al., 2020*a*)—and April

1 through April 4 (the sample in the random encouragement design). Using a point estimate for *λ*_0_ and bounding Pr(*I* = 1|*E* = 0) resulted in tighter bounds: [0.891,0.938] for the earlier dates and [0.806,0.905] for the later ones. NUHI eligibility guidelines were broadened after March 19 (see Section 2), so the results from early April lead to lower estimates of the undetected rate. Recall that the estimate for *λ*_0_ is the peak weekly case-load during the H1N1 outbreak in the fall of 2009. Since *τ*_*U*_ is a strictly decreasing function of *λ*_0_, it follows that a smaller value of *λ*_0_ shifts the upper bound on the identified set upwards, so there is some conservatism to this value. Because *λ*_0_ also enters the lower bound for *P* (*I* = 1|*E* = 0) in a monotonically increasing way, the relationship between *λ*_0_ and *τ*_*L*_ is nonlinear. To clarify the sensitivity of our estimates to *λ*_0_, we plot our bounds as a function of *λ*_0_ in the Appendix Section B. These estimates are qualitatively robust to a range of plausible values for *λ*_0_.

#### Random screening method

We estimate Pr(*I* = 1|*E* = 1) over the same dates when deCODE tested the random sample, i.e. April 1 through 4. We estimate the lower bound of *θ* is about 68%. Adding Assumption 2 to address the exclusion of quarantined individuals barely changes the lower bound, while decreasing the upper bound substantially (by about 7 ppt). The decrease in the upper bound is driven by the monotonicity assumption on the infection rate for those with *Q* = 0, *Y* = 0. The width of the bounds in both cases reflects in the worst-case Pr(*I* = 1|*Q* = 0, *Y* = 0) = 0 and the fact that quarantined individuals were excluded from participation. The results suggest that randomized encouragement designs have limited identifying power if one wishes to avoid uncomfortable assumptions about the sub-population that do not comply with the randomization. Consequently, public health officials should design direct randomization methods that sample from the *entire* population in order to gain more informative estimates of pandemic parameters.

## 6 Conclusion

This paper seeks to estimate the fraction of COVID-19 infections that were undetected by strict eligibility criterion for testing (the ‘undetected rate’) in the early stages of the pandemic. Using data from Iceland, we conclude that the undetected rate was very high: our preferred estimates of the lower bound imply that at least 80% of infected individuals were ineligible for medical testing, even after the medical system broadened its criteria. The undetected rate will be higher than estimates of the fraction of infections that are asymptomatic (Mizumoto et al., 2020; Nishiura et al., 2020), since ineligible individuals can be mildly symptomatic. The results continue to have implications for the United States and other countries, especially as phased re-opening plans are executed.

Our methods carry some caveats. We rely on several key assumptions on the eligible share ofthe population and medical system testing. Moreover, we do not consider the presence of false positives and false negatives. We believe that assuming zero false positives is reasonable with the testing methods used here, but simple thought experiments show that this is not innocuous: even a false positive rate of 0.5% (with no false negatives) could explain the infection rate we observe among the ineligible and imply a true undetected rate of 0%. Our paper serves as a stepping stone and subsequent analysis that relaxes these assumptions is an important undertaking for econometricians.

## Data Availability

All data are publicly available. We use counts of tests and confirmed COVID-19 infections from the Icelandic Health Directorate's COVID-19 information website. We also use counts of cases reporting flu-like symptoms from the influenza website of the country's Health Directorate. We use quarantine numbers from the Department of Civil Defense's announcement page.

https://www.landlaeknir.is/servlet/file/store93/item32967/influensulik.einkenni.uppfaersla.a.vef.okt2017.AG.xls_%C3%A1n%20myndar%20og%20aldursdreifingu.xls

https://www.covid.is/data

https://www.almannavarnir.is/utgefid-efni/

## 7 Acknowledgements

The development of the paper has benefited from numerous discussions. In particular, we thank Isaiah Andrews, Eric Budish, Kevin Chen, Charles Manski, Francesca Molinari, Rami Tabri, and Elie Tamer. We also are grateful to Guðrún Aspelund, Thor Aspelund, Bergdís Bjoörk Sigurjoónsdóttir, Sigríður Haraldsdóttir, and Agnes Gísladóttir for their insight into the Icelandic COVID-19 testing efforts and data. Aspelund acknowledges support by the National Science Foundation Graduate Research Fellowship under Grant No. 1122374. All errors are our own.

## A Proofs

### A.1 Identified Sets

We start by proving the propositions related to the random screening identification.

#### *Proof of Proposition 1*.

The proof follows by the direct method. First note that *θ* = 1−Pr(*E* = 1|*I* = 1) by the complement rule. Next, Bayes rule and the law of total probability yields

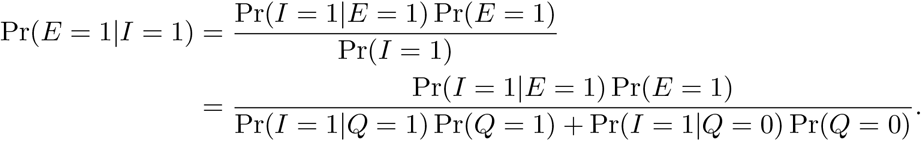

To complete the proof, we need to consider the denominator. In accordance with the text, Pr(*I* = 1|*Q* = 1) *∈* [0, 1] and Pr(*I* = 1|*Q* = 0) *∈* [*L*_0_, *U*_0_] where

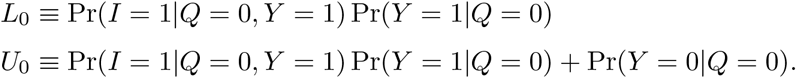

Consequently,

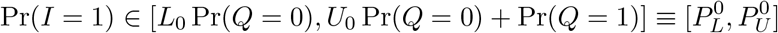

and immediately

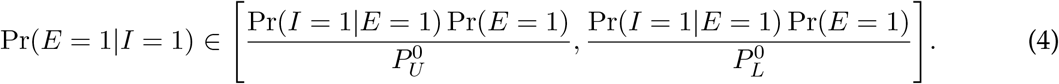

Apply the complement rule to (4) to complete the proof.

□

*Proof of Proposition 2*. The proof follows the same steps as that of Proposition 1. The only difference is that now 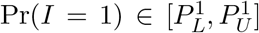 under Assumption 2 and Pr(*I* = 1|*Q* = 0) *∈* [*L*_0_, *U*_1_], where 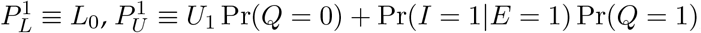, and *U*_1_ = Pr(*I* = 1|*Y* = 1, *Q* = 0).

□

Now we turn to the odds ratio approach. We start with the alternative representation of *θ*, which is simply an algebraic result.

**Lemma A**.**1**. *An alternative representation of θ is*

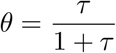

*Where*

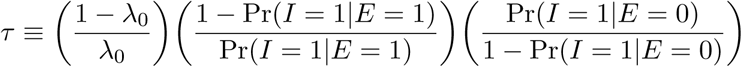

*and λ*_0_ ≡ Pr(*E* = 1|*I* = 0).

*Proof*. The proof proceeds by the direct method. By Bayes rule and the law of total probability,

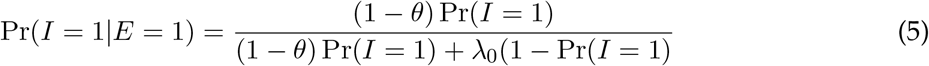

And

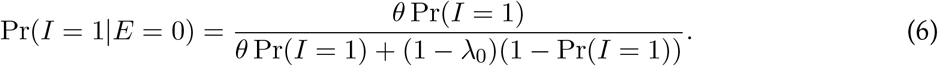

Take the ratio of odds ratios implied by (5) and (6), we have

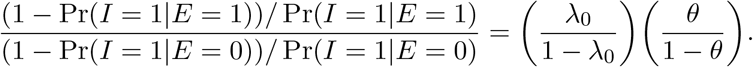

Rearranging,

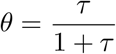

□

as required.

Now that we have established the representation, we prove Proposition 3.

*Proof of Proposition 3*. The proof proceeds by the direct method. We first derive bounds for Pr(*I* = 1|*E* = 0). To this end, we note that

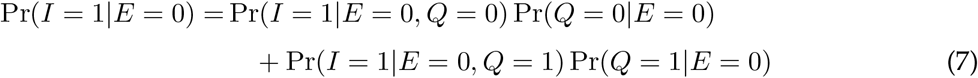

by the law of total probability.

We consider each term in (7) individually. Bayes law and the fact that Pr(*E* = 0|*Q* = 1) *∈* [0, 1] yields that Pr(*Q* = 1|*E* = 0) *∈* [0, Pr(*Q* = 1)*/* Pr(*E* = 0)]. Immediately, the complement rule allows us to conclude Pr(*Q* = 0|*E* = 0) *∈* [1 − Pr(*Q* = 1)*/* Pr(*E* = 0), 1]. By the law of total probability,

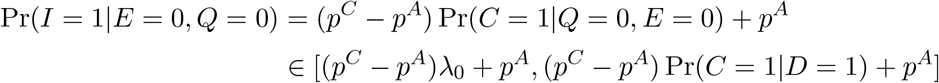

where the bounds hold by Part 1 of Assumption 3. Finally, Pr(*I* = 1|*E* = 0, *Q* = 1) *∈* [0, Pr(*I* = 1|*T* = 1)] by Part 2 of Assumption 3. Putting these all together, we have bounds for Pr(*I* = 1|*E* = 0) given by

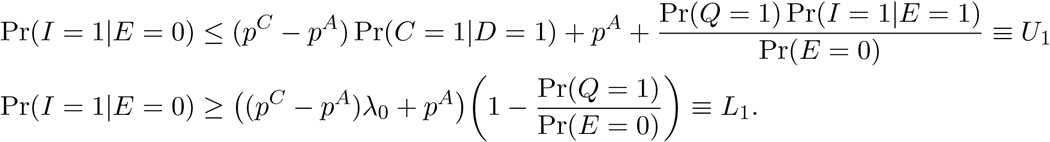

Evaluate *τ* at *U*_1_ and *L*_1_ and the result follows from the fact that 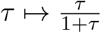 is strictly increasing over [0, 1].

□

## Confidence Set Assumptions and Validity

**Assumption 4**. *𝒫 satisfies*

1. *There exists ε*_*∗*_, *ε*^*∗*^ *∈* (0, *∞*) *such that* 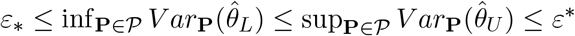
2. *The following weak convergence holds for all subsequences* 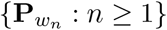 *in P as n* → *∞:*

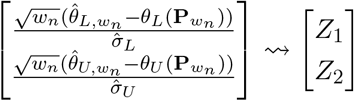

*where* 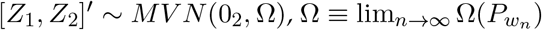, *and* 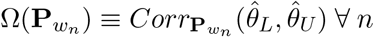.

Part 2 is a uniform asymptotic normality assumption that allows one to establish the validity of *CS*_*n*_(1 − *α*) and Part 1 ensures that the elements of 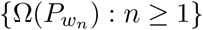are well-defined. Since data in the random screening is based on an i.i.d. sample of the Icelandic population and the random variables are almost-surely bounded, such assumption is reasonable.

**Proposition 4**. *Under Assumption 4, CS*_*n*_(1 − *α*) *satisfies*

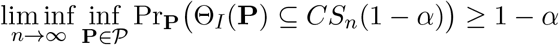

*Proof*. It suffices to show that for an arbitrary sequence *{***P**_*n*_ : *n* ≥ 1*}* in *P*,

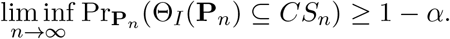

The limit inferior is smallest subsequential limit, so trivially there exists a subsequence 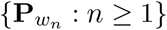 of *{***P**_*n*_ : *n* ≥ 1*}* such that

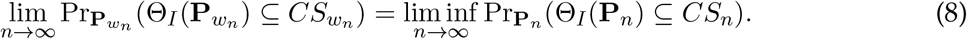

By definition of 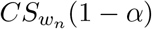, it follows that

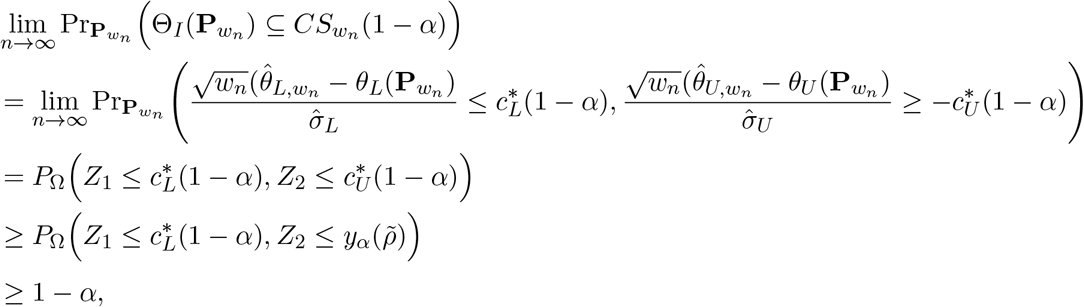

Where 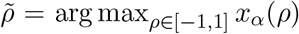. The last equality holds by Assumption 4 plus the symmetry of the standard normal distribution, the first inequality holds because 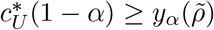, and the last inequality holds by construction of the critical value (i.e. 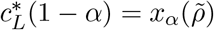). Combining this result with (8) completes the proof.

□

## B Sensitivity to *λ*_0_ in Odds Ratio Approach

The odds ratio approach to identifying *θ* requires the parameter *λ*_0_ = Pr(*E* = 1|*I* = 0), which we interpret to be the underlying rate of cold/flu symptoms in the general public. As we describe in Section 5.1, we use Directorate of Health weekly influenza count data from 2009 through early 2017 to estimate *λ*_0_ as the greatest one-week fraction of Icelanders reporting flu symptoms at health clinics, which is 0.62%.

To clarify whether our estimates from the odds ratio approach are sensitive to *λ*_0_, Figure 1 plots the estimated identified regions for *θ* as a function of *λ*_0_. The blue shaded area corresponds to odds ratio estimates using testing data from March 13-19, and the red shaded area corresponds to odds ratio estimates using testing data from April 1-4. The dashed vertical line denotes the calibrated value of *λ*_0_ used in our baseline estimates. As this figure indicates, our estimates are reasonably robust to a range of values for *λ*_0_.

**Figure 1:**
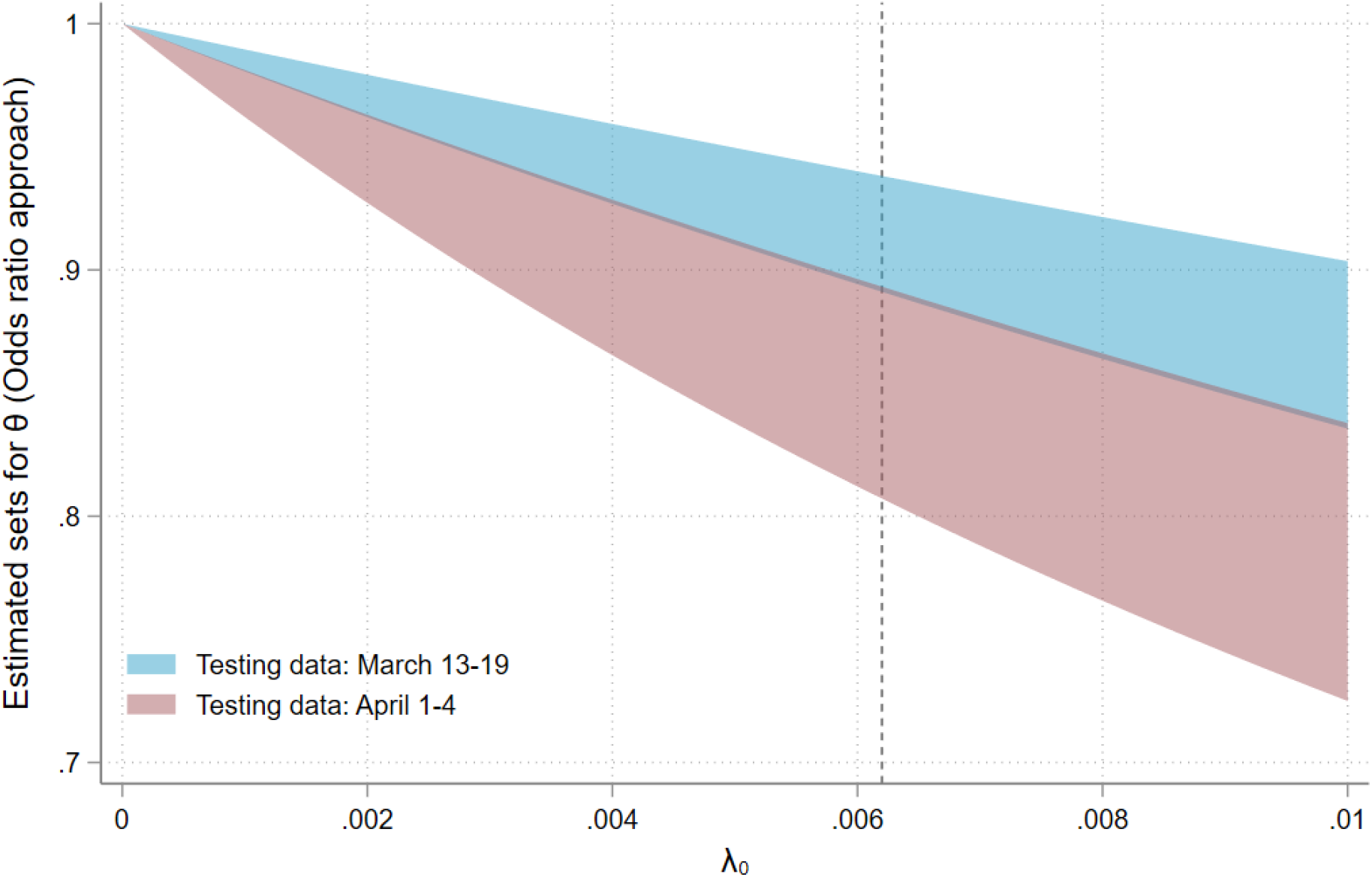
Sensitivity of Odds Ratio Estimates to *λ*_0_. *Notes*: This figure plots estimates of *θ* using the odds ratio approach outlined in section 3.2 as a function of the parameter *λ* _0_. The dashed line indicates the value of *λ* _0_ used in our baseline estimates, presented in Table 2. The blue shaded region corresponds to our estimates using testing data from March 13-19, and the red shaded region corresponds to our estimates using testing data from April 1-4.

## C Manski and Molinari (2020) for Iceland

As noted in Remark 1, we implement Equation (13) from Manski and Molinari (2020), using the publicly available data from Iceland to bound the daily population infection rate. In this section, we follow their notation: on date *d, C*_*d*_ is an indicator for infection, *T*_*d*_ for having been tested, and *R*_*d*_ for have a positive test result. In particular, we aggregate counts of infections and tests from deCODE and NUHI for each day, available at https://www.covid.is/data. We take as the population of Iceland 364,134, as in our main analysis (see Appendix Section D). We use Manski and Molinari‘s upper and lower bounds on the false negative rate *P* (*C*_*d*_ = 1|*T*_*d*_ = 1, *R*_*d*_ = 0) *∈* [0.1, 0.4]. The method requires taking the minimum and maximum of values before and after the given date *d*. We take these values from the first date of double-digit positive test count (March 4) to the day before the positive test count returned to single digits for the first time (April 12). We then report the bounds for the dates relevant to our analysis, March 13 through April 4, in Table 3.

## D Data Construction

See the text for definitions of the various terms. This section describes the construction of each parameter in detail. Table 1 summarizes the values.

- Population: Statistics Iceland estimates that the population of Iceland was 364,134 on January 1,2020.^14^
- Pr(*E* = 1): Sum the total count of NUHI tests over the relevant dates, to match the deCODE testing dates, and divide by the Icelandic population. The daily counts are available at https://www.covid.is/data.
- Pr(*I* = 1|*E* = 1): Calculate the total count of NUHI infected cases and divide by the total count of NUHI tests for the relevant dates, to match the deCODE testing dates. The daily counts are available at https://www.covid.is/data.
- *λ*_0_: In considering the proportion of non-infected who are eligible for medical system testing, we seek reasonable estimates of the number of individuals not infected with COVID-19 but with serious flu-like symptoms. We use Directorate of Health weekly influenza count data from 2009 through early 2017 to estimate *λ*_0_ by the greatest one-week fraction of Icelanders reporting flu symptoms at health clinics.^15^ This period occurred in November 2009 during the H1N1 pandemic and was 0.63%.
- *p*^*C*^, *p*^*A*^: Divide infection count by the test count for symptomatic and asymptomatic individuals, respectively, from Table 1 of Guðbjartsson et al. (2020*a*) for March 13 through March 19 (columns 4 and 5) and Table 1 of Guðbjartsson et al. (2020*b*) for April 1 through April 4 (columns 8 and 9).
- Pr(*C* = 1|*D* = 1): Divide the test count on symptomatic individuals by the total test count from Table 1 of Guðbjartsson et al. (2020*a*) for March 13 through March 19 (columns 4 and 5)
- and Table 1 of Guðbjartsson et al. (2020*b*) for April 1 through April 4 (columns 8 and 9).
- Pr(*Q* = 1): The COVID-19 website only reports the most updated count of individuals under quarantine, but we require the proportion quarantined at the time of testing. We use the number of quarantined individuals on March 19, the last day of deCODE testing reported in Guðbjartsson et al. (2020*b*) from the report of the Department of Civil Defense on March 20 and on April 1, the first day of testing of the sample from the randomized encouragement design (since deCODE randomized over the population in the days preceding April 1).^16^
- Pr(*Y* = 1|*Q* = 0): deCODE invited a random sample of Icelanders for testing, so the quarantine rate for the invitees will be the same as the population quarantine rate. Therefore, multiply the total number of invitees in the randomized encouragement design (6,782) by the proportion of the population not quarantined on April 1. This is the number of non-quarantined invitees. Divide the total number of invitees who show up for testing (2,283) by the (calculated) number of non-quarantined invitees. This gives the compliance rate of those not quarantined. The number of invitees and participants are reported in Guðbjartsson et al. (2020*b*).
- Pr(*I* = 1|*Y* = 1, *Q* = 0): Divide the number of infected individuals in the random screening sample (reported in column 9 of Guðbjartsson et al. (2020*b*)) by the total number of participants in the random screening (reported in column 8 of Guðbjartsson et al. (2020*b*)).

1 The Centers for Disease Control and Prevention (CDC) in the United States recommend these testing criteria. See https://www.cdc.gov/coronavirus/2019-nCoV/hcp/clinical-criteria.html.

2 See publicly available case counts at https://www.covid.is/data.

3 A broad treatment of partial identification is presented in Tamer (2010).

4 https://www.landlaeknir.is/um-embaettid/greinar/grein/item39194/Skilgreind-ahaettusvaedi---Defined-high-risk-areas

5 Quarantines last 14 days or until the individual develop symptoms and are tested positive for SARS-CoV-2 by NUHI. Individuals with a positive test, whether quarantined or not at the time of testing, must enter a 10-day isolation with more stringent restrictions than those for quarantine. See https://www.covid.is/categories/how-does-isolation-work and https://www.covid.is/categories/how-does-quarantine-work.

6 Just as with NUHI results, one could calculate these rates from daily infection and test counts from deCODE available on https://www.covid.is/data.

7 From this discussion, it is clear that we assume a complete partition: no NUHI-eligible individuals report for deCODE testing. We believe this assumption is reasonable, since deCODE testing was targeted to asymptomatic individuals and those who had never been tested in the medical system. See, for example, the Health Directorate’s description of the deCODE testing program at https://www.landlaeknir.is/um-embaettid/greinar/grein/item38808/Questions-and-answers-regarding-novel-coronavirus-in-China.

8 See survey descriptions in the weekly reports at https://www.landlaeknir.is/influensa/. The Directorate compiled numbers from these weekly reports in a spreadsheet. See Section 5.1 and Appendix Section D.

9 For instance, see https://www.fda.gov/media/136151/download for information on analytical estimates of sensitivity/specificity of an RT-PCR test in use in the United States. Another resource aggregating analytical estimates can be found at https://www.finddx.org/covid-19/sarscov2-eval-molecular/molecular-eval-results/.

10 The data is insufficient to compute some joint probability masses that are inputs to the covariances.

11 The variances implied by the delta methods are continuous functions of the nonidentified parameters, so Weierstrass’ theorem implies that the maximum exists.

12 English: https://www.covid.is/data

13 Only in Icelandic: https://www.landlaeknir.is/servlet/file/store93/item32967/influensulik.einkenni.uppfaersla.a.vef.okt2017.AG.xls_%C3%A1n%20myndar%20og%20aldursdreifingu.xls

14 https://px.hagstofa.is/pxen/pxweb/en/Ibuar/Ibuarmannfjoldi1_yfirlityfirlit_mannfjolda/MAN00000.px/table/tableViewLayout1/?rxid=cdf68733-56cd-41c8-8801-d73ca7ded1cd

15 In Icelandic: https://www.landlaeknir.is/servlet/file/store93/item32967/influensulik.einkenni.uppfaersla.a.vef.okt2017.AG.xls_%C3%A1n%20myndar%20og%20aldursdreifingu.xls

16 March 20, in Icelandic: https://www.almannavarnir.is/utgefid-efni/stoduskyrsla-koronaveira-covid-19-20032020/?wpdmdl=24791 April 1, in Icelandic: https://www.almannavarnir.is/utgefid-efni/stoduskyrsla-koronaveira-02032020-2/?wpdmdl=24580

## References

Boston Public Health Commission. 2020. “Results Released for Antibody and COVID-19 Testing of Boston Residents.” Press Release, May 18, 2020.

Bullard, Jared, et al. 2020. “Predicting infectious SARS-CoV-2 from diagnostic samples.” Clinical Infectious Diseases.

Cocci, Matthew D, and Mikkel Plagborg-Møller. 2019. “Standard errors for calibrated parameters.” Working Paper.

Guðbjartsson, Daníel F, et al. 2020a. “Early Spread of SARS-CoV-2 in the Icelandic population.” MedRxiv Pre-Print: https://doi.org/10.1101/2020.03.26.20044446.

Guðbjartsson, Daníel F., et al. 2020b. “Spread of SARS-CoV-2 in the Icelandic population.” New England Journal of Medicine.

Horowitz, Joel L, and Charles F Manski. 2000. “Nonparametric analysis of randomized experiments with missing covariate and outcome data.” Journal of the American statistical Association, 95(449): 77–84.

Indiana State Dept. of Health. 2020. “COVID-19 Random Sample Study: Preliminary Results.” Presentation, May 13.

Li, Ruiyun, et al. 2020. “Substantial undocumented infection facilitates the rapid dissemination of novel coronavirus (SARS-CoV2).” Science, 368: 489–493.

Manski, Charles F., and Francesca Molinari. 2020. “Estimating the COVID-19 infection rate: Anatomy of an inference problem.” Journal of Econometrics.

Mizumoto, Kenji, Katsushi Kagaya, Alexander Zarebski, and Gerardo Chowell. 2020. “Estimating the asymptomatic proportion of coronavirus disease 2019 (COVID-19) cases on board the Diamond Princess cruise ship, Yokohama, Japan, 2020.” Euro Surveillance, 25.

New York Governor’s Press Office. 2020. “Amid Ongoing COVID-19 Pandemic, Governor Cuomo Announces Results of Completed Antibody Testing Study of 15,000 People Showing 12.3 Percent of Population Has COVID-19 Antibodies.” Press Release, May 2, 2020.

Nishiura, Hiroshi, et al. 2020. “Estimation of the asymptomatic ratio of novel coronavirus infections (COVID-19).” International Journal of Infectious Diseases, 94.

Office for National Statistics. 2020. “Coronavirus (COVID-19) Infection Survey pilot: 5 June 2020.” Statistical Bulletin, June 5.

Public Health Agency of Sweden. 2020. “Fö rsta resultaten frå n på gå ende undersö kning av antikroppar fö r covid-19-virus.” Report, May 20, 2020.

Public Health England. 2020. “Weekly Coronavirus Disease 2019 (COVID-19) Surveillance Report.” Report, Week 22.

Qiu, Jane. 2020. “Covert coronavirus infections could be seeding new outbreaks.” Nature News.

Ramdas, Kamalini, Ara Darzi, and Sanjay Jain. 2020. “Test, re-test, re-test’: using inaccurate tests to greatly increase the accuracy of COVID-19 testing.” Nature Medicine.

Russell, Timothy W., et al. 2020. “Using a delay-adjusted case fatality ratio to estimate underreporting.” Centre for Mathematical Modeling of Infectious Diseases Repository.

Spanish Ministry of Health. 2020. “Estudio nacional de sero-epidemilogía de la infección por SARS-CoV-2 en España: Informe preliminar 13 de Mayo de 2020.” Report, May 13, 2020.

Streeck, Hendrick, Gunther Hartmann, Martin Exner, and Matthias Schmid. 2020. “Vorläufiges Ergebnis und Schlussfolgerungen der COVID-19 Case-ClusterStudy (Gemeinde Gangelt).” Report.

Tamer, Elie. 2010. “Partial Identification in Econometrics.” Annual Review of Economics, 2: 167–195.

Vogel, Gretchen. 2020. “Antibody surveys suggesting vast undercount of coronavirus infections may be unreliable.” Science News.

Wu, Joseph T., et al. 2020a. “Estimating clinical severity of COVID-19 from the transmission dynamics in Wuhan, China.” Nature Medicine, 26: 506–510.

Wu, Xiaodong, Bo Fu, Lang Chen, and Yong Feng. 2020b. “Serological tests facilitate identification of asymptomatic SARS-CoV-2 infection in Wuhan, China.” Journal of Medical Virology.

Zheng, Shufa, et al. 2020. “Viral load dynamics and disease severity in patients infected with SARS-CoV-2 in Zhejiang province, China, January-March 2020: retrospective cohort study.” BMJ, 369.

